# Periodic Testing and Estimation of STD-HIV Association

**DOI:** 10.1101/19002345

**Authors:** B.R. Mâsse, P. Guibord, M.-C. Boily, M. Alary

## Abstract

**Background:** The validity of measures used in follow-up studies to estimate the magnitude of the HIV-STD association will be the focus of this paper. A recent simulation study by Boily *et al* [1] based on a model of HIV and STD transmission showed that the relative risk (*RR*), estimated by the hazard rate ratio (*HRR*) obtained by the Cox model had poor validity, either in absence or in presence of a real association between HIV and STD. The *HRR* tends to underestimate the true magnitude of a non-null association. These results were obtained from simulated follow-up studies where HIV was periodicaly tested every three months and every month for the STD.

**Aims and Methods:** This paper extends the above results by investigating the impact of using different periodic testing intervals on the validity of *HRR* estimates. Issues regarding the definition of exposure to STDs in this context are explored. A stochastic model for the transmission of HIV and other STDs is used to simulate follow-up studies with different periodic testing intervals. *HRR* estimates obtained with the Cox model with a time-dependent STD exposure covariate are compared to the true magnitude of the HIV-STD association. In addition, real data are reanalysed using the STD exposure definition described in this paper. The data from Laga *et al* [2] are used for this purpose.

**Results:** (1) Simulated data: independently of the magnitude of the true association, we observed a greater reduction of the bias when increasing the frequency of HIV testing than that of the STD testing. (2) Real data: The STD exposure definition can create substantial differences in the estimation of the HIV-STD association. Laga *et al* [2] have found a *HRR* of 2.5 (1.1 - 6.4) for the association between HIV and genital ulcer disease compared to an estimate of 3.5 (1.5 - 8.3) with our improved definition of exposure.

**Conclusions:** Results on the simulated data have an important impact on the design of field studies. For instance when choosing between two designs; one where both HIV and STD are screened every 3 months versus one where HIV and STD are screened every 3 months and monthly, respectively. The latter design is more expensive and involves more complicated logistics. Furthermore, this increment in cost may not be justified considering the relatively small gain in terms of validity and variability.

## Introduction

### Issues in the study of the HIV-STD interaction

Evidence to support the role of STDs as cofactors in HIV transmission has been growing in recent years. It is believed that STD prevalence is an important factor possibly explaining heterogeneities and epidemiological patterns of HIV infection across countries and between different regions within countries [2, 3, 4, 5]. Results of many epidemiological studies have suggested an association between HIV infection and past or present infection with different STDs. Despite international consensus of the possible link between STD and HIV infection, estimates of the magnitude of association vary greatly between STDs, risk groups, countries, study designs [2, 6, 7, 8, 9, 10].

The objective of such epidemiological studies is to ascertain whether the presence of a STD in the HIV-susceptible person increases the susceptibility to HIV infection. The detection of a causal relationship between STDs and HIV, even under a simple mode of action, is made problematic by many issues such as the diagnosis of HIV infection, the measurement of exposure to STDs (whether the STD was present at the time of HIV infection), the presence of confounding behavioral variables (e.g., condom use) and the different study designs. The assessment of independent risk factors is often made problematic by the presence of confounding variables such as multiple sex partners, receptive anal sex or differential exposure to infections due to a preferencial choice of sexual partners. With retrospective studies there is often a problem in determining the precise sequence of events of STD and HIV infection. A history of STD infection does not imply that a STD was present at the time of, or before, the acquisition of HIV infection. But even in follow-up studies, the exact time of HIV infection is difficult to determine because (1) HIV and STD testing is usually performed at periodic intervals (e.g., every 3 or 6 months) and (2) the HIV undetectable period. Testing of HIV and STD at periodic intervals will create interval-censored data which occurs when the exact time of infection is unknown but the infection is known to have occurred within some time interval. For example, if an individual is tested for HIV every six months and seroconverts at the second visit (i.e. at 12 months), then we will only know that he has seroconverted between the sixth and twelfth month of follow-up. The same situation applies with STD testing. However with HIV, the determination of the time interval is complicated by the undetectable period (usually not a problem with STDs). Specifically, the HIV undetectable period corresponds to the period of time between HIV infection and detectable HIV antibodies (i.e. seroconversion) in an individual. This period varies according to the screening test used and between individuals. Therefore, the HIV undetectable period has to be accounted for in order to determine if an individual was exposed to a STD at the time of HIV infection. For the above example and given a HIV undetectable period of 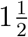 to 3 months; the possible time period of infection with HIV is between 3 and 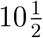 months. Estimation of the association between HIV infection and other STDs needs to consider the fact that the time of infection is interval-censored and that exposition to STD is time-dependent. Effectively, the STD status of individuals followed during the study changes with time due to infection (or reinfection) and recovery. Complication comes from the fact that these two issues need to be addressed simultaneously in the statistical analysis.

### Objectives

In this paper, we assess the extent to which the periodic testing of STD and HIV and the definition of exposure influence the estimates of the magnitude of the HIV-STD association. As explained earlier, the definition of exposure to a STD in a follow-up study is somewhat difficult when analyzing data where HIV and STD are tested at periodic intervals. There is no unique way of defining this exposure and several definitions can be used. It is however important to realize that a different definition can have a different impact, yet difficult to assess clearly, on the results in terms of bias. In many field studies the choice of the frequency of testing is often arbitrary, mostly based on the costs of testing and field logistics.

The relative risk (*RR*) defines the true degree to which HIV transmission is enhanced by the STD cofactor. The hazard rate ratio (*HRR*) is used to estimate the *RR* and detect the association between HIV infection and STD cofactor (e.g., gonorrhoea). The results presented in this paper are based on data simulated with a proportional hazard stochastic model of HIV-l/STD transmission in a sexually heterogeneous population [1], Monte-Carlo simulations were used to generate population-based data recording the history of each individual under different assumptions concerning the magnitude of the enhancement of HIV transmission by the STD. Using this database, various assumptions can be formulated to represent variations in the design of follow-up studies.

Boily and Anderson [1] have previously shown that the *HRR* tends to underestimate the true magnitude of the HIV-STD association based on STD testing every month and HIV testing every 3 months. Their STD exposure definition was the one used by Laga *et al*. [2] with a HIV undetectable period of 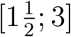 months. Laga’s definition of STD exposure basically adds 4 months of STD exposure to the observed STD exposure. For example, if a subject is STD positive at month 1, 2, and 3 and STD negative at the other visits; his STD exposure for the purpose of analysis is defined as STD positive at month 1, 2, 3, 4, 5, 6, and 7 and STD negative at the other visits. In this paper, Laga’s definition is refined and the impact of different periodic interval of testing is investigated. These issues are studied using simulated and real data (from a field study). The exact time of infection and time interval based on different frequencies of HIV and STD testing were used to assess their impact on the estimates of the *RR*. In addition, a discussion and recommendations for study planning and data analysis are provided.

## Methods

### Stochastic model for the transmission of HIV

The stochastic model used for the simulations can be viewed as a continuous, non-homogeneous time Poisson process with four disease states representing the different stages of HIV and STD infection (details of which can be found in Boily and Anderson [1]). In the absence of HIV, the process describes an open but stable population. The sexually active population is stratified by gender and sexual activity (based on the rate of sexual partner acquisition). The number of individuals per disease state at time *t* is represented by 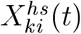, where *k* = 1 if female; *k* = 2 if male; *i* = 1,…, 6 for the different sexual activity classes; *s* = 0 if susceptible to STD, *s* = 1 if diagnosed with a current STD; *h* = 0 if susceptible to HIV and *h* = 1 if infected with HIV. *Figure 1* represents the flow chart of the principal interactions between the key variables and shows the flow of individuals from one disease state to another. In this model, the transition between states (e.g., the possible events) occurs at different times according to the random process determined by the probabilities of the model. These probabilities are the product of the population size and the hazard rate at which a given event occurs in a fixed population at fixed time (e.g., the net rate of HIV infection of the susceptible population is defined by 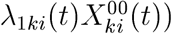. The per capita rates of HIV infection is given by λ_1*ki*_(*t*) in the susceptibles while that of the STD infected is given by

**Figure 1:**
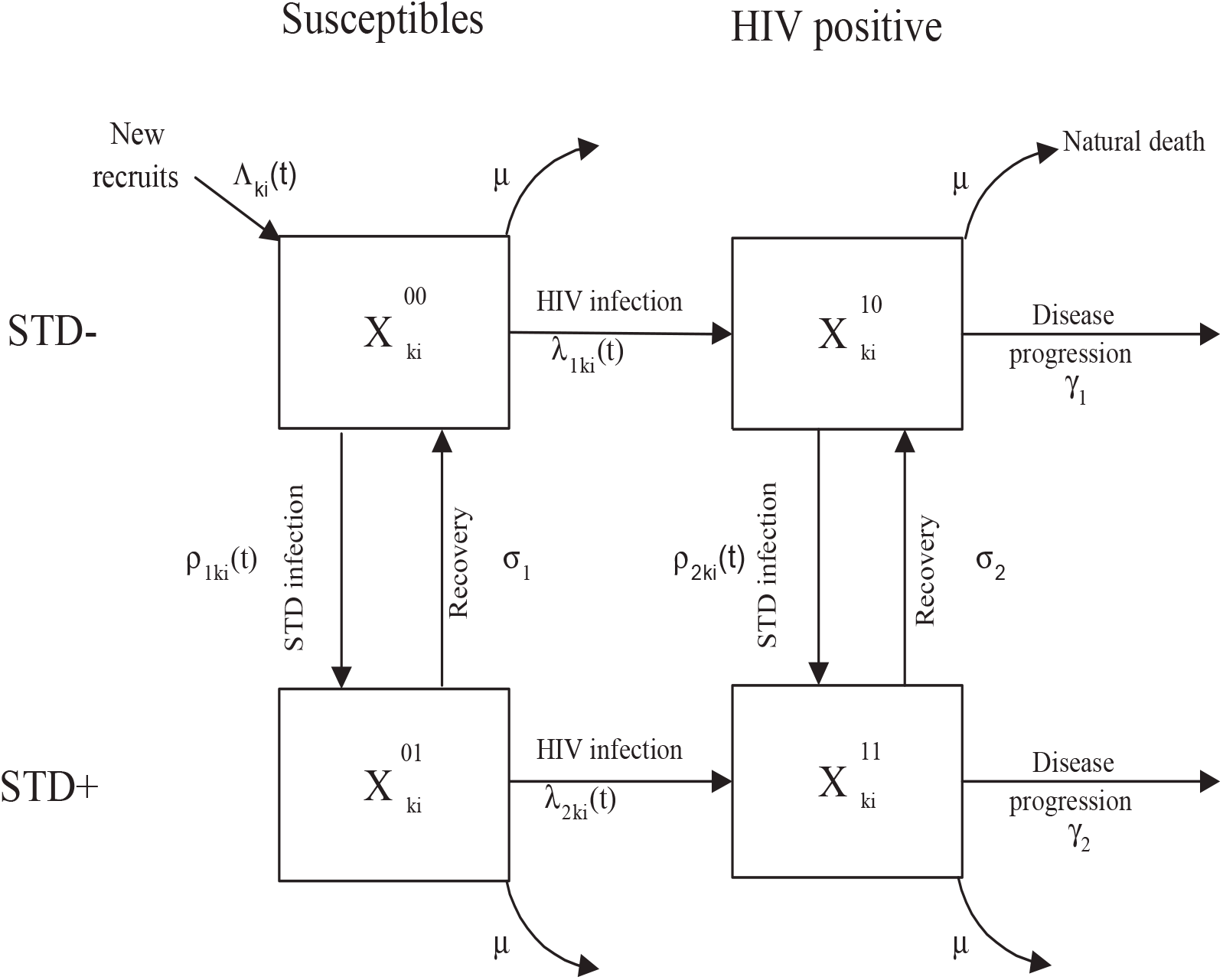
Compartimental model representing the flow of individuals from one disease state to another and the relationships between the key population variables : those susceptible to HIV and to the cofactor STD, 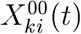; those susceptible to HIV and infected with the cofactor STD, 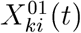; those infected (asymptomatic) with HIV and with the cofactor STD, 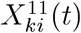; those infected (asymptomatic) with HIV and susceptible to the cofactor STD, 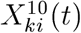. The transition rates are : *μ* for the natural death; Λ_*ki*_(*t*) for the birth rate; *ρ*_1*ki*_(*t*) and *ρ*_2*ki*_(*t*) for the infection with the STD in HIV- and HIV+ individuals respectively; *λ*_1*ki*_(*t*) and *λ*_2*ki*_(*t*) for the infection with the HIV in STD- and STD+ individuals respectively; *σ*_1_ and *σ*_2_ for the recovery from the STD in HIV- and HIV+ individuals respectively; *γ*_1_ and *γ*_2_ for the progression to AIDS in HIV- and HIV+ individuals respectively. Here *k* = 1, 2 for the gender; *i* = 1,…, 6 for the different sexual activity classes.

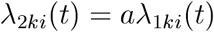

and we can define the relative risk by

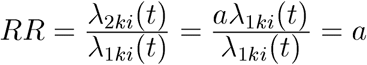

The latter equation shows that the *RR* is equal to the HIV transmission enhancement factor *a*, who defines the degree to which HIV transmission is enhanced by the STD cofactor. The resulting risk is proportional, thus supporting the proportional hazard assumption underlying the hazard rate ratio (*HRR*) obtained by the Cox model. The absence of an association (*RR*=1) and the presence of an association (*RR*=4) will be studied in this paper. Further details for the parameter values used in the simulations and a complete definition of the net rates can be found in Boily and Anderson [1].

### Definition of exposure to STD

The quality of the estimates of the association is highly dependent on the definition of exposure. However, the definition of exposure to a cofactor that is (1) time-dependent, (2) periodically screened and (3) influenced by the HIV undetectable period is problematic. In order to circumvent difficulties created by the periodic testing, it is common practice to use the midpoint of the interval between a negative and a positive test result as the exact time of infection. The midpoint represents the mean of the distribution which is assumed to be uniform in the interval. Therefore we assume that seroconversion occurs at the midpoint between the last negative HIV serological test result and the first positive one. Similarly, STD infection (recovery) are assumed to have occurred at the midpoint between a positive (negative) test result and the last negative (positive) visit. Note that the STD recovery time may be set to be a certain number of weeks after a positive test when it is known that an appropriate treatment was initiated immediately after the positive visit.

In this paper, the STD exposure definition of a given individual is allowed to vary over time and the HIV undetectable period is assumed to be 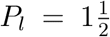 to *P*_*u*_ = 3 months. Individual *i* will be considered exposed for an additional period of exposure, say *A*_*i*_ months, following a positive diagnosis of STD where *A*_*i*_ will depend on the HIV undetectable period (say [*P*_*l*_;*P*_*u*_]) and the length of the HIV testing interval (say *I*_*i*_). Specifically, we define *A*_*i*_ as:

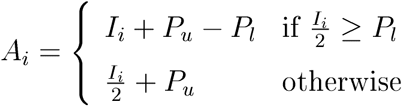

Note that the length of the interval *I*_*i*_ can vary between individuals. This permits different HIV periodic testing between individuals where, for example, some individuals may be tested for HIV every month and other every 3 months. However, this is rarely the case in practice. Therefore, *I*_*i*_ is held constant (and so is *A*_*i*_) leading to 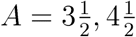, and 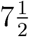 for a HIV undetectable period of 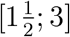 months and a length of HIV testing interval *I* of 1, 3, and 6 months, respectively. At this point we need to define the time of infection and recovery (in months) from a STD. Let 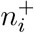 be the number of infection time for individual *i* where the infection time is noted as 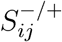 and 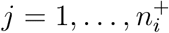. Similarly, we can define the STD recovery time by 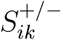 where 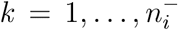 with 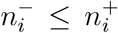. Note that 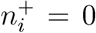 and 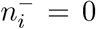 if individual *i* was negative at every visit (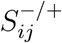 and 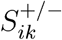 are not defined for 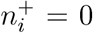 and 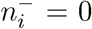). The observed STD exposure from an individual could be represented by the sequence 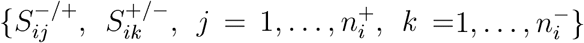. The additional exposure *A* is then added to the 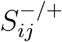 ’s which yields the augmented sequence 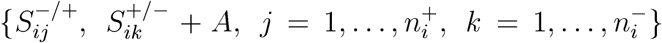. Note that the augmented sequence is used only for the analysis on the HIV-STD association. It would be inappropriate to use the augmented sequence to estimate association between STDs and possible risk factors.

The following example illustrates the determination of the augmented sequence with 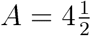. Let the sequence

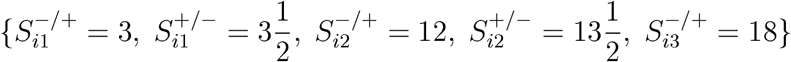

represents individual *i* who was infected at 3 months and recovered at 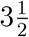, was reinfected at 12 months and recovered at 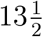 months, and was finally re-infected at 18 months without recovering. His observed STD exposure intervals are: 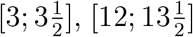, and [18; ∞) (note that here ∞ represents the end of the study). With the addition of 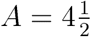 the augmented sequence is

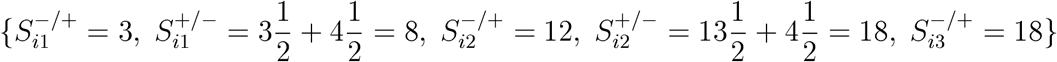

which yields the STD exposure intervals (for the purpose of analysis): [3; 8] and [12; ∞). This definition is not entirely satisfactory since an interval where an HIV infection could occur is not covered when the HIV testing interval *I* is larger than 2*P*_*l*_. Let 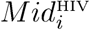 be the midpoint estimate of the time of HIV infection for individual *i*, then the above definition does not cover the interval 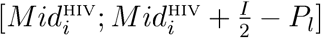. For example, if HIV is tested every 6 months (*I* = 6 months and 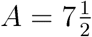 months) and individual *i* was HIV positive at 18 months. The interval [15; 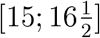] is not covered in the case where this individual has a STD exposure sequence of 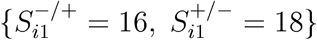 which yields the augmented sequence 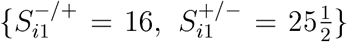. Thus, this individual is considered not exposed at 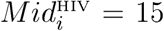 months eventhough a HIV infection could have occured between 16 and 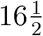 months while this individual was STD positive during that period.

The definition is refined by substracting the excess *E* to the infection time 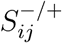. The definition of *E* and A are then:

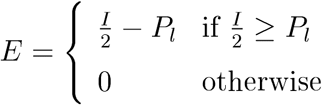

and

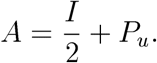

The augmented sequence of STD exposure becomes 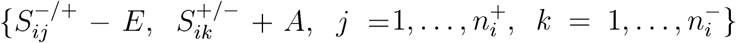. For the above example and the refined definition, the augmented sequence with 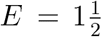 and *A* = 6 is 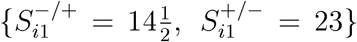 where this individual is now considered exposed at 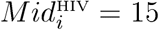 months. *Figures 2A* and *2B* graphically illustrate two examples of the above STD exposure definition for a periodic testing of HIV of 3 and 6 months, respectively.

**Figure 2:**
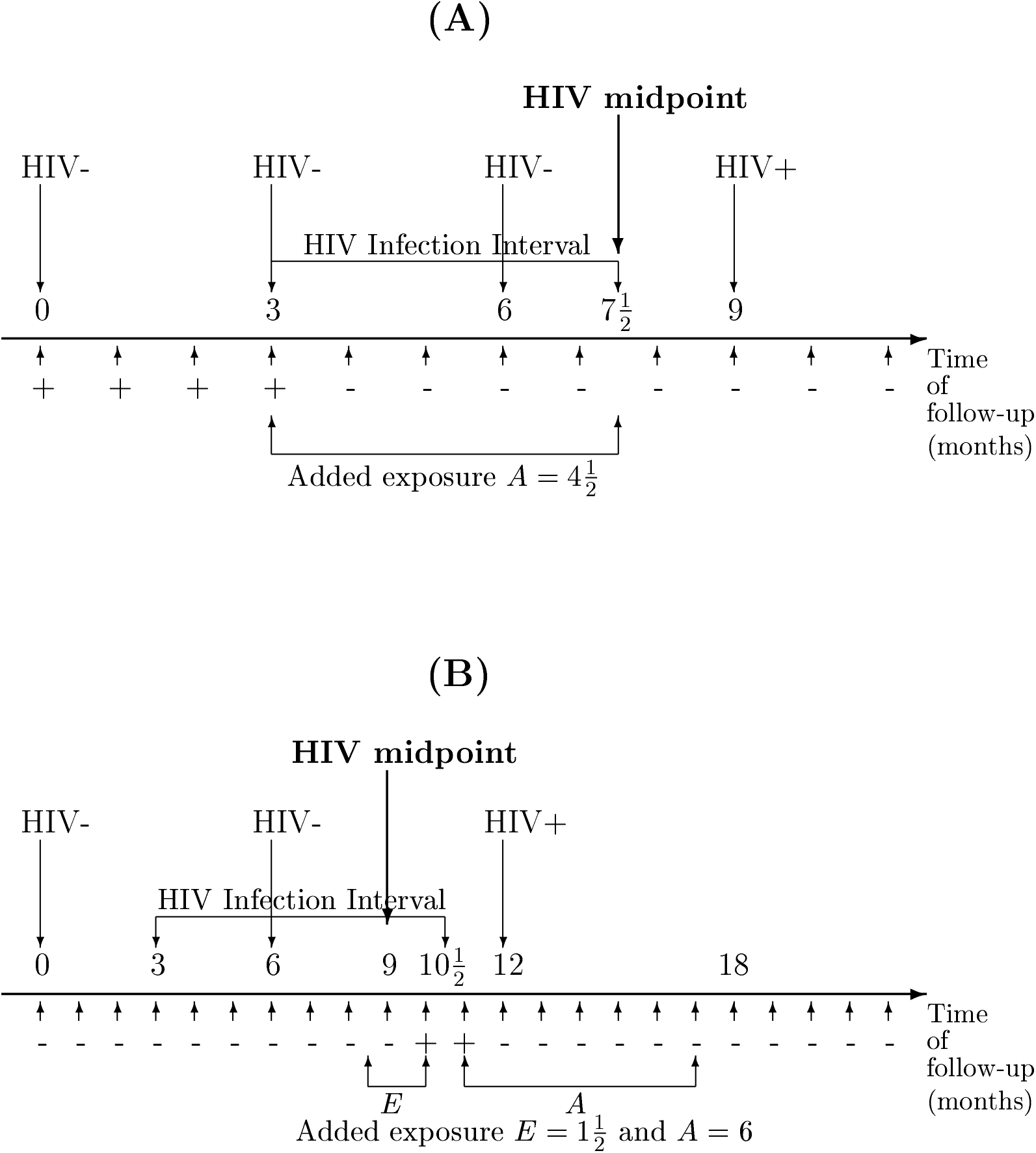
**(A)** This figure illustrates the STD exposure of individual *i* with an observed STD exposure sequence of 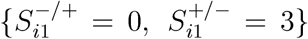 where HIV (↓) is tested every 3 months and every month for the STD (↑). The resulting augmented sequence, for the purpose of analysis, is 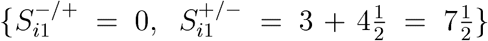. **(B)** This figure illustrates the STD exposure of individual *i* with an observed STD exposure sequence of 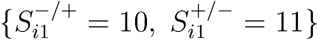 where HIV (↓) is tested every 6 months and every month for the STD (↑). The resulting augmented sequence, for the purpose of analysis, is 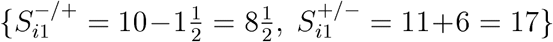. In both cases **(A)** and **(B)**, the individual will be considered exposed at the time 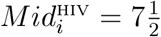 and 9 months, respectively.

This definiton is somewhat different than the one used by Laga *et al*. [2, 6] where HIV was tested every 3 months and monthly for the STDs. Using the same HIV undetectable period, they have used *A* = 4 months for the augmented sequence where our definition yields 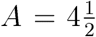 months. The difference lies on the dependence of Laga’s definition on the length of the STD testing interval. Our definition relies only on the length of the HIV infection interval. The difference can be seen with the help of *Figure 2A*. Laga *et al*. are considering that the individual will recovered at 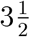 and that they only need to set *A* = 4 months in order to expose this individual to STD at 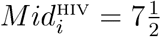. The HIV infection interval should not depend on the length of the STD testing interval. Let say that an individual receives an appropriate treatment for his STD during 10 days starting at 3 months. His recovery time is less than 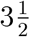 and he will not be considered exposed at 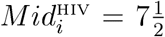 if *A* = 4 months is used. Clearly, HIV infection could have occured while he was still STD positive. This reason led us to use the length of the HIV infection interval for the determination of *A* and *E*, in fact, this length equals *A* + *E*. The difference between these two definitions of exposure can create substantial differences in the estimates of *RR* as it will be seen in the *Results* section when we reanalyse Laga’s data with our definition.

### Data analysis

A Cox proportional hazard model with a time-dependent covariate was used to estimate the *HRR* adjusted for sexual activity. The time-dependency is treated as a binary covariate indicating the STD status of the individual at different time intervals. This type of analysis can be handled using the model based on the counting process method [11, 12, 13] which has several advantages: it has the ability to easily code time-dependent covariate and it is usually more efficient than the other strategy [14, 15], The counting process method is now available in SAS release 6.11 [16] and integrated in S-PLUS release 3.4 [17], In addition, the pooled logistic model was used where intervals of equal length are treated as short follow-up studies. At the beginning of each interval of observation, the STD status is recorded and considered as a potential predictor of outcome in the interval. With this model, it is assumed that the time of events is not known exactly but occurs during a fixed interval of follow-up. Afterwards, the information contained in the intervals are then pooled into a single sample for the analysis [18, 19, 20],

### Simulations

Prospective follow-up studies were designed as follows. For each gender, at the beginning of the epidemic, a random sample of 1200 HIV-negative individuals is chosen among the four highest sexual activity classes. This cohort of patients is then followed for a fixed period of time (e.g., 1, 3 or 5 years). At the end of each study, the *HRR* is estimated, and a new random sample is chosen from those still susceptible to HIV in the population. The new sample is then followed for a further period of time. In this manner, the magnitude of the association is estimated every time a study is terminated during the course of the HIV epidemic. Samples of the high activity classes were chosen because most studies conducted in the field are carried out among female commercial sex worker (CSW) or STD clinic patients. During the follow-up period, no individual joins or leaves the cohort except as a result of death. The validity of the measures of association between STD and HIV is assessed by comparing the estimated *HRR* with the true magnitude of the association *RR*. Fifty repetitions of 20 years are simulated.

A FORTRAN program was written to perform the simulations for a large population and at the beginning of each stochastic simulations parameters values were specified and initial conditions defined [1], It is important to note that parameter values were chosen to represent a population typical of Sub-Saharan Africa, where most of the field studies on HIV-STD association had taken place. Values for parameters and initial value conditions that, on average, characterize the typical course of STD and HIV infections are estimated from published studies [1], The analysis was performed under the assumption that the real magnitude of the association between HIV and the presence of cofactor STD under the alternative hypothesis that the cofactor STD enhances HIV transmission by a factor of 4 (*RR*=4), or does not enhance HIV transmission (*RR*=1). The STD cofactor was assumed to be gonorrhea because its natural history is relatively well documented rendering the selection of parameter values feasible. The beginning of the epidemic was initiated by the introduction of two seropositive females, one in each of the fifth and sixth sexual activity class.

Follow-up studies with varying frequencies of screening for HIV and STD are simulated. HIV testing at every month, 3 and 6 months and STD testing at every 2 weeks, month, and 3 months are investigated. Note that the pair 2 weeks for STD and 6 months for HIV was not simulated because it was judged unrealistic in held studies along with the pair 3 months for STD and monthly for HIV which implies a frequency of screening for HIV higher than the STD frequency, which is neither common in held studies.

### Real data

Previously published data on a cohort of prostitutes in Zaire [2] were analyzed to evaluate the impact of the different exposure definitions on the analysis of real data. Our aim with this complementary analysis is to see if the definition of exposure used in this article would change the results obtained previously by Laga *et al*. [2]. The study reports the results of an intensive clinic-based intervention including condom promotion as well as monthly STD testing, testing for HIV every 3 months and treatment of female CSW. The study population consists of women from Kinshasa who identified themselves as CSW. This population has a high STD prevalence, a 35% HIV-1 prevalence, and a high level of risky behaviors. Consenting HIV-1 seronegative women were enrolled in a cohort study and followed for up to 3 years. Details of the study can be found in Laga *et al* [2].

## Results

This section presents the results obtained from the Cox model analyses of the simulation data. It was interesting to evaluate the pooled logistic model, which accounts for the grouped nature of the data, in order to investigate if it will give better results in the context of simultaneous HIV and STD testing. In our simulations, this method gave similar results than with the Cox model for all testing scenarios and length of follow-up. Therefore, only the results based on the Cox model are presented.

### Analysis with the exact time of events

The objective of the exact time analysis is to validate the results generated by the model without the impact of the midpoint imputation and exposure definition. The endpoint considered was the exact time of HIV and STD infections and recovery from STD. As expected, the results of *Table 1A* shows that *HRR* estimates based on exact times are unbiased. It can also be seen that results from the female cohort are less variable than the male cohort bacause of the higher overall sexual activity that characterize the females of this population. As a result, more females get infected in the course of the epidemic. As we would expect, the estimates become less variable with the increasing duration of the follow-up.

**Table 1:** **(A)** *HRR* estimates (adjusted for sexual activity) obtained from a Cox model with a time-dependent STD exposure covariable when the exact time of events are used. Medians (inter-quartile ranges) of the *HRR* estimates obtained over the simulated data are presented in the table. **(B)** *HRR* estimates (adjusted for sexual activity) obtained from a Cox model with a time-dependent STD exposure covariable when different periodic testing are used in studies with 3 years of follow-up. Medians (inter-quartile ranges) of the *HRR* estimates obtained over the simulated data are presented in the table.

| Exact time of events |  |  |  |  |
| --- | --- | --- | --- | --- |
| Length of follow-up | True $RR = 1$ | | True $RR = 4$ | |
|  | Females | Males | Females | Males |
| 1 year | 1.00 (0.36) | 1.09 (1.12) | 3.97 (1.07) | 4.19 (2.92) |
| 3 years | 0.97 (0.27) | 1.03 (0.73) | 4.03 (0.77) | 4.07 (1.96) |
| 5 years | 0.99 (0.25) | 1.03 (0.62) | 3.95 (0.76) | 4.15 (1.85) |

(B)
| Periodic testing of STD and HIV |  |  |  |  |  |
| --- | --- | --- | --- | --- | --- |
| Frequency of testing | | True $RR = 1$ | | True $RR = 4$ | |
|  |  | Females | Males | Females | Males |
| STD | HIV |  |  |  |  |
| 2 weeks | 1 month | 0.87 (0.20) | 0.95 (0.64) | 2.24 (0.41) | 2.44 (1.21) |
| 1 month | 1 month | 0.86 (0.22) | 0.97 (0.62) | 2.30 (0.43) | 2.56 (1.21) |
| 2 weeks | 3 months | 0.84 (0.20) | 0.92 (0.53) | 2.05 (0.39) | 2.19 (1.12) |
| 1 month | 3 months | 0.84 (0.21) | 0.92 (0.56) | 2.04 (0.43) | 2.21 (1.07) |
| 3 months | 3 months | 0.75 (0.19) | 0.83 (0.68) | 1.85 (0.43) | 2.24 (1.29) |
| 1 month | 6 months | 0.77 (0.20) | 0.87 (0.50) | 1.70 (0.35) | 1.81 (0.92) |
| 3 months | 6 months | 0.68 (0.16) | 0.77 (0.55) | 1.45 (0.44) | 1.74 (1.01) |

### Periodic testing of STD and HIV

*Table 1B* shows the adjusted *HRR* estimated from studies with 3 years of follow-up, for a true *RR*=*1* and 4, for males and females, and for different periodic testing. For *RR*=*1, HRR* underestimates slightly the true *RR*. However for *RR*=4, a substantial under-estimation of the true *RR* is observed for all testing scenarios. The *HRR* are about half the size of the true *RR* (2 instead of 4) for all testing scenarios. Decreasing the frequency of testing of STD or HIV increases the (negative) bias. More importantly, the frequency of testing for HIV has a greater impact on the estimates than the frequency of STD testing. This can be seen by comparing the results for the monthly testing of STD and HIV testing at every 1, 3 and 6 months. It is clear that increasing the frequency of HIV testing reduces the bias. On the other hand, if we compared the results of HIV testing every 3 months and STD testing every 2 weeks, 1 month and 3 months, we can see that increasing the testing frequency for STD does not substantially reduce the bias. These simulations indicate that there seems to be a small gain, in terms of reducing the bias, of testing for STD more often than the testing for HIV. Therefore, if possible the design should be chosen preferably on the basis of the frequency of HIV testing, not on the frequency of STD screening. The same conclusions are derived from studies with 1 and 5 years of follow-up for both *RR*=1 and *RR*=4 (results not shown).

In terms of costs, the results have a substantial implication. Suppose that a subject is enrolled in a 3 years study with monthly STD testing and testing of HIV at every 3 months (this is a frequency of testing often encountered in practice). Then, 37 visits will be required to complete the study (assuming a baseline testing and that testing of STD and HIV can be done in one visit). If the STD is screened every 3 months, the number of visits drops down to 13. Although there is a slight gain in terms of validity (i.e. reduction of bias) for monthly STD testing compared to 3 months, the substantial reduction of the total number of visits greatly outweigh the gain in validity. For the above example (and assuming no loss to follow-up), if 100 subjects are enrolled, 3700 visits (37 visits × 100 subjects) compared to 1300 visits (13 visits × 100 subjects) will be required. If one is willing to sacrifice precision in order to increase validity, a study with 70 subjects and HIV and STD testing every 2 months would generate 1330 visits (19 visits × 70 subjects) is a good alternative. Field logistics are also simplified when both testing of STD and HIV are performed at the same visit.

### Analysis of real data

Comparisons between the analyses with different exposure definitions are given in *Table 2*. Small differences are observed in the *HRR* estimates for well defined time-independent cofactors such as age and duration of prostitution. However for more problematic time-dependent STD cofactors such as gonorrhea, chlamydia, trichomoniasis and genital ulcer disease, the differences are more important. The largest difference is observed with genital ulcer disease where the *HRR* estimate increases from 2.5 to 3.5. The significant results of most of the *HRR* estimates of this study are in agreement with the enhanced effect of STDs in HIV infection found in the literature [1, 2, 4, 6, 7, 9, 10], Furthermore, *HRR* estimates based on our definition are generally larger than those of Laga *et al* [2], Given the results of the simulations, this suggests that the bias is probably smaller and that the magnitude of the STD-HIV association is probably larger than what was first found by in Laga *et al* [2].

**Table 2:** Results obtained from a multivariate analysis using a Cox model with timedependent covariables on the data of Laga *et al* [2], The *HRR* with the corresponding 95% *CI* was computed using the STD exposure definition of Laga *et al* [2] and the one described in this paper.

| Factors | Definition of exposure used |  |
| --- | --- | --- |
|  | Laga *et al* | In this paper |
| **Time-independent** |  |  |
| Age <20 years | 1.5 (0.7-3.2) | 1.7 (0.8-3.7) |
| 20-29 years | 1.4 (0.8-2.6) | 1.5 (0.8-2.8) |
| +30 years | 1 | 1 |
| Duration of prostitution | 1.5 (0.9-2.4) | 1.5 (0.8-2.4) |
| **Time-dependent** |  |  |
| Irregular condom use | 1.6 (1.0-2.6) | 1.7 (1.1-2.8) |
| Gonorrhoea | 2.3 (1.4-3.7) | 1.9 (1.1-3.2) |
| Chlamydial infection | 0.9 (0.5-1.6) | 1.2 (0.8-2.1) |
| Trichomoniasis | 1.7 (1.1-2.8) | 2.2 (1.4-3.6) |
| Genital ulcer disease | 2.5 (1.1-6.4) | 3.5 (1.5-8.3) |

## Discussion

A good testing schedule should be one that provides valid estimates and does not result in unjustified costs and heavy logistics. We observed that the frequency of STD testing is not as important as the frequency of HIV testing on the validity of *HRR* estimates. Indeed, what we observed for all follow-up durations is a larger bias when we decreased the frequency of HIV testing than when we decreased the frequency of STD testing. The choice of the periodic testing interval is crucial when considering the importance of the costs and logistics associated in a follow-up study. For example, a STD testing frequency every 2 weeks will be more costly but will not reduce substantially the negative bias in the estimates of the association compared to a monthly testing, irrespectively of the HIV testing frequency. In this case, the costs and logistics in terms of number of visits required outweigh the reduction in the *HRR* bias. Thus, it is better to allocate resources to increase the frequency of HIV testing than the frequency of STD testing.

New diagnostic tests are currently being developed. These tests will serve to shorten the HIV window of detection. This new kind of tests efficiently detects the level of p24 antigens [21, 22] or the level of DNA (or RNA) viral load [23, 24, 25] by polymerase chain reaction (PGR) instead of only the level of antibodies. The p24 test have the drawback to be less sensitive than the DNA test. The addition of these tests for antibody testing would decrease the window period for detecting individuals recently infected with HIV. This window period corresponding to the period between HIV infection and detection, is actually around 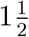 to 3 months. It could be reduced to 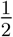 to 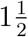 months with the addition of these new tests [26], Until now the impact of these new tests on the estimates of the HIV-STD association remains unknown. By decreasing the window period, this would reduce the bias and therefore would increase the validity. The importance of the effect is to be determine and further simulations should be done in order to ascertain the effect of these new tests. Better tests would serve to ameliorate diagnosis but they are too costly to be used in the held at the moment. Thus, actual screening tests for HIV and STD would still be used in most studies.

The method used in this paper to deal with the interval-censored data, the definition of the STD exposure, and the HIV undetectable period is somewhat *ad-hoc*. Statistical models applied to right censored data are not, unfortunately, readily applicable to interval-censored data. More research should be devoted on developing methods for analyzing interval-censored data that handles time-dependent cofactors. Few computationally intensive and not yet widely implemented methods are currently available [27, 28], However, most current methods cannot take into account time-dependent cofactors and interval-censored data simultaneously. Recent works on logspline models render possible the analysis of data containing both interval-censored data and time-dependent covariates [29]. Unfortunately, the numerical implementation of this method is still in development (should be available in a futur S-PLUS release).

The general conclusions of the study can apply to any STD (e.g., gonorrhoea) if we make the assumption that the association between HIV and the presence of cofactor STD is enhanced by a certain factor (e.g., 4) in a female or male population. The simulations presented in this paper focus on females CSW and males from STD clinics because of their relatively high incidences of STD and HIV in developing countries and that most published studies focus on these high activity groups.

## Data Availability

The data that support the findings of this study are available from Laga et al [2]. Restrictions apply to the availability of these data, which were used under licence for this study. Data are available from Marie Laga.

## Acknowledgments

MCB thanks the NHRDP for financial support. MA is supported by the NHRDP and FRSQ. The authors would like to thank Marie Laga for kindly providing the data from the study in Zaire.

